# Detecting and isolating false negatives of SARS-CoV-2 primers and probe sets among the Japanese Population: A laboratory testing methodology and study

**DOI:** 10.1101/2020.10.07.20208264

**Authors:** Wataru Tsutae, Wirawit Chaochaisit, Hideyuki Aoshima, Chiharu Ida, Shino Miyakawa, Hiroko Sekine, Afzal Sheikh, Iri Sato Baran, Toshiharu Furukawa, Akihiro Sekine

## Abstract

**Objectives:** In this study, a comparative study between primers from Japan’s and US’s disease control centers was conducted. As further investigation, virus sequence alignment with primers’ oligonucleotide was analyzed.

**Design or methods:** 11,652 samples from Japanese population were tested for SARS-CoV-2 positive using recommended RT-PCR primer-probe sets from Japan National Institute of Infectious Disease (NIID) and US Centers for Disease Control and Prevention (CDC).

**Results:** Of the 102 positive samples, 17 samples (16.7% of total positives) showed inconsistent results when tested simultaneously for the following primers: JPN-N2, JPN-N1, CDC-N1, and CDC-N2. As a result, CDC recommended primer-probe sets showed relatively higher sensitivity and accuracy. Further virus sequence alignment analysis showed evidences for virus mutation happening at primer’s binding sites.

**Conclusions:** The inconsistency in the RT-PCR results for JPN-N1, JPN-N2, CDC-N1, and CDC-N2 primer-probe sets could be attributed to differences in virus mutation at primers’ binding site as observed in sequence analysis. The use of JPN-N2 combined with CDC-N2 primer produces the most effective result to reduce false negatives in Japan region. In addition, adding CDC-N1 will also help to detect false negatives.

## INTRODUCTION

The global COVID-19 pandemic has spread across various continents in diverse methods and speed while opening up discussion for technological and scientific questions pertaining methodology of testing accuracy among diverse viral strains.

On the issue of testing sensitivity and accuracy, RT-PCR has been the gold standard testing method for SARS-CoV-2 as opposed to other rapid testing methods (Corman et al. 2020; Wu et al. 2020). However, each country’s infectious disease authority has established its own primer-probe sets guidelines and protocols, thereby, the global testing community lacks a “Standardized Universal Primer(s)” (SUP) that is foolproof for the COVID-19 patients among various populations today. As a result, RT-PCR testing accuracy and results may vary depending on which primer was used, most likely resulting in false negative and/or false positive calls associated with RT-PCR testing.

There are two commonly known factors associated with inaccurate testing results for COVID-19 using RT-PCR testing. First is the failure to retrieve sufficient amounts of viral RNA typically associated with the timing of when the sample is administered or the method of how a sample is collected. For example, a nasopharyngeal swab may be unable to obtain sufficient amounts of RNA if it does not come in contact at a nasal position where the presence of the virus is concentrated. In the case of saliva collection kit, use during the first several days of viral contact may result in insufficient amounts of RNA.

For both nasopharyngeal swab and saliva kit, low amounts of RNA occurring at later stage among discharged or recovering patients has tendency to show extremely low RNA count. While many countries have issued a standard 14-day quarantine, Genesis Healthcare, a licensed clinical laboratory in Tokyo, Japan, has confirmed through tested samples where the recovering and discharged patient is still testing positive after 14 day despite low RNA count. Though this confirmation does not necessarily imply that the patient is infectious after 14 days of quarantine as observed in a study (Cheng et al. 2020), while the test result remains positive, it supports recent study which outlines the long duration of the RNA-positive tail and calls for reconsideration of containment strategy (Mina et al. 2020).

The second factor is the lack of a standardized international testing method for COVID-19 using a set of common primer(s) among different nations/populations or hereby referred to as SUP. Different primers used by different country’s testing protocols prevent effective tracking of pandemic due to differences in false positives and/or false negative results due to viral genetic variation which may have been introduced from different regions.

Researchers has started to recognize this problem as more data about the viral genomes become available. In particular, a researcher group evaluated five assay panels for possible loss of sensitivity due to genetic variability of the virus (Peñarrubia et al. 2020). Their conclusion was in accordance with what has been observed in our study.

In this paper, the RT-PCR result discrepancy for SARS-CoV-2 testing is summarized and compared among different primers recommended by NIID and CDC. Further, virus mutation as a plausible cause of discrepancy is investigated and discussed.

## METHODS

### Study Design

Japan’s COVID-19 RT-PCR public health observation is unique in that while Wuhan, China was locked down from January 23, 2020 till April 8^th^, 2020, Japan’s border was still open to remainder of China until March 9^th^, 2020 and to US and Europe until March 26, 2020. This time gap in closing the international border leaves an inquiry that COVID-19 could have possibility entered Japan at different times from both East and West.

On March 11^th^ and March 19^th^, NIID announced the testing protocols for COVID-19 testing by outlining two primer-probe sets JPN-N2 and JPN-N1 targeting nucleocapsid region, N of SARS-CoV-2 as shown in Figure 1.

**Figure 1.**
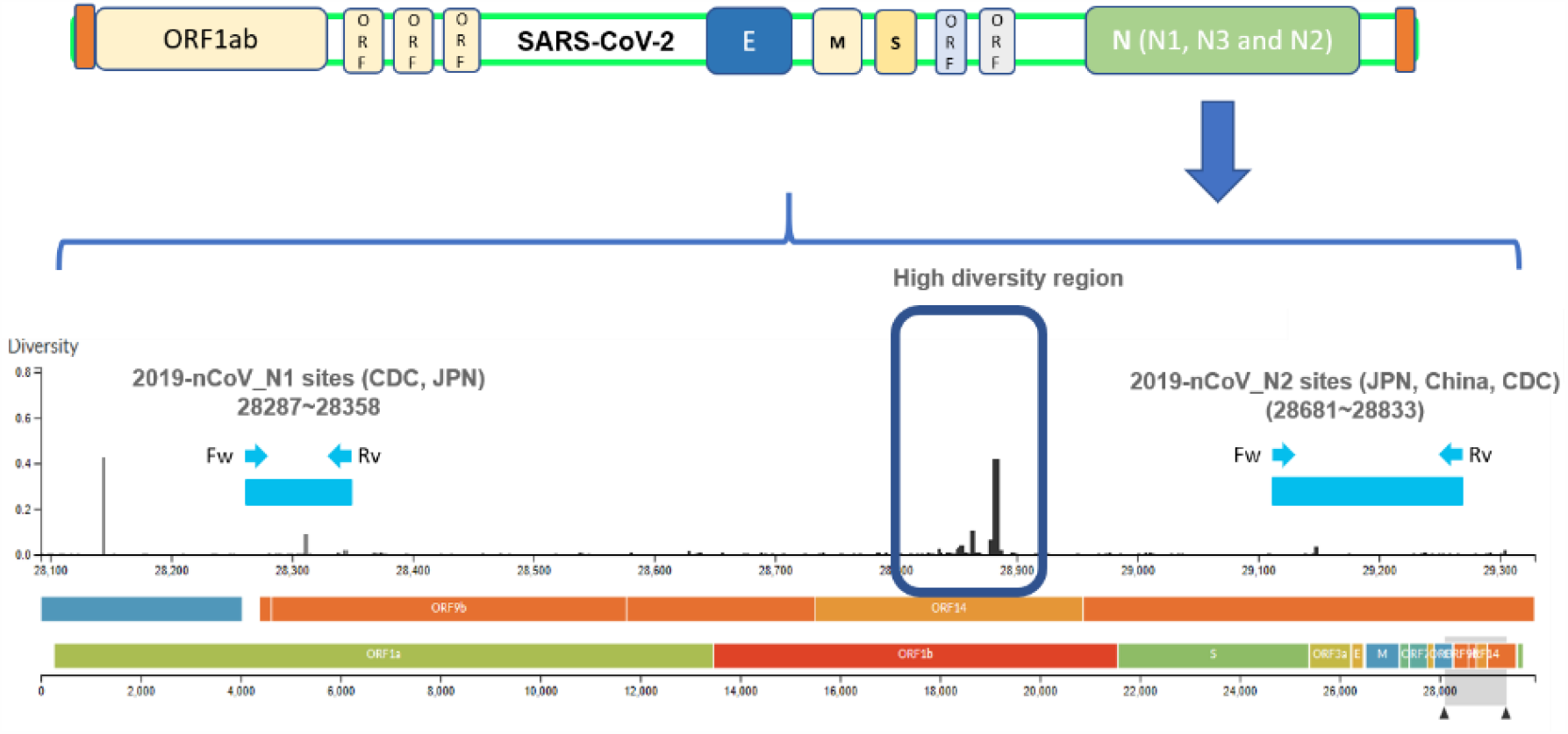
Structural representation of SARS-CoV-2 and primer/probe sites. The diversity sites were sourced from Hadfield et al. (2018).

JPN-N2 has seen consistently higher primer sensitivity than JPN-N1. As a result, it is sought that NIID later eliminated JPN-N1 to only reflect JPN-N2 in calling positive samples.

Due to the continuing dialogue by the medical community to retest samples whose results were not considered to reflect patient’s symptoms, testing JPN-N1 was continued and CDC-N1 and CDC-N2, both of which are located in nucleocapsid region N of SARS-CoV-2, were added to identify whether JPN-N2 primer was adequate to identify COVID-19 positive cases for all samples.

### Sample Collection

All COVID-19 positive testing samples were obtained between April, 2020 and August, 2020 and were either obtained from saliva or nasopharyngeal test kits. The nasopharyngeal testing included two swabs per person and were administered to maximize virus collection to rule out miscalling due to low RNA count. Both nasopharyngeal and saliva samples were immediately immersed in RNA preservation solution after extraction to inactivate the virus while maintaining RNA stability during transport. Samples were delivered to the Genesis Healthcare’s PCR testing facility within 24 hours at temperatures between 20° and 27° Celsius. All samples’ RNA extraction was immediately conducted followed by RT-PCR test. The entire RNA extraction to RT-PCR testing process was completed within 6 hours of receipt at the same laboratory using the below testing method.

### COVID-19 RNA Collection Method, Extraction and RT-PCR Testing Method

Saliva samples were collected using saliva RNA sample collection kit by Zeesan Biotech Co., Ltd. Nasopharyngeal swab samples were collected using virus RNA sample collection kit by Zeesan Biotech Co., Ltd. Saliva samples were added 0.5 ml 20% DTT to remove viscosity, vortex and incubate at 50° Celsius for 10 minutes, then centrifuged at 1,500 rpm for 10 minutes.

180 μl of supernatant of DTT-treatment saliva samples and 180 μl of nasopharyngeal swab suspension were used for RNA extraction. RNA extraction was performed by MGISP-960 system (MGI Tech Co., Ltd). RT-qPCR for specific amplification of the N gene or E gene of SARS-CoV-2 was performed using LightCycler480 system?(Roche Diagnostics K.K.). Final reaction volume was 12.5 μl, including 2.0 μl of RNA template, 6.25 μl of One Step PrimeScript III RT-qPCR Mix (TAKARA BIO INC.), and forward primer, reverse primer and probe (primer and probe sequence, final concentration was shown in Table 1.

**Table 1.**
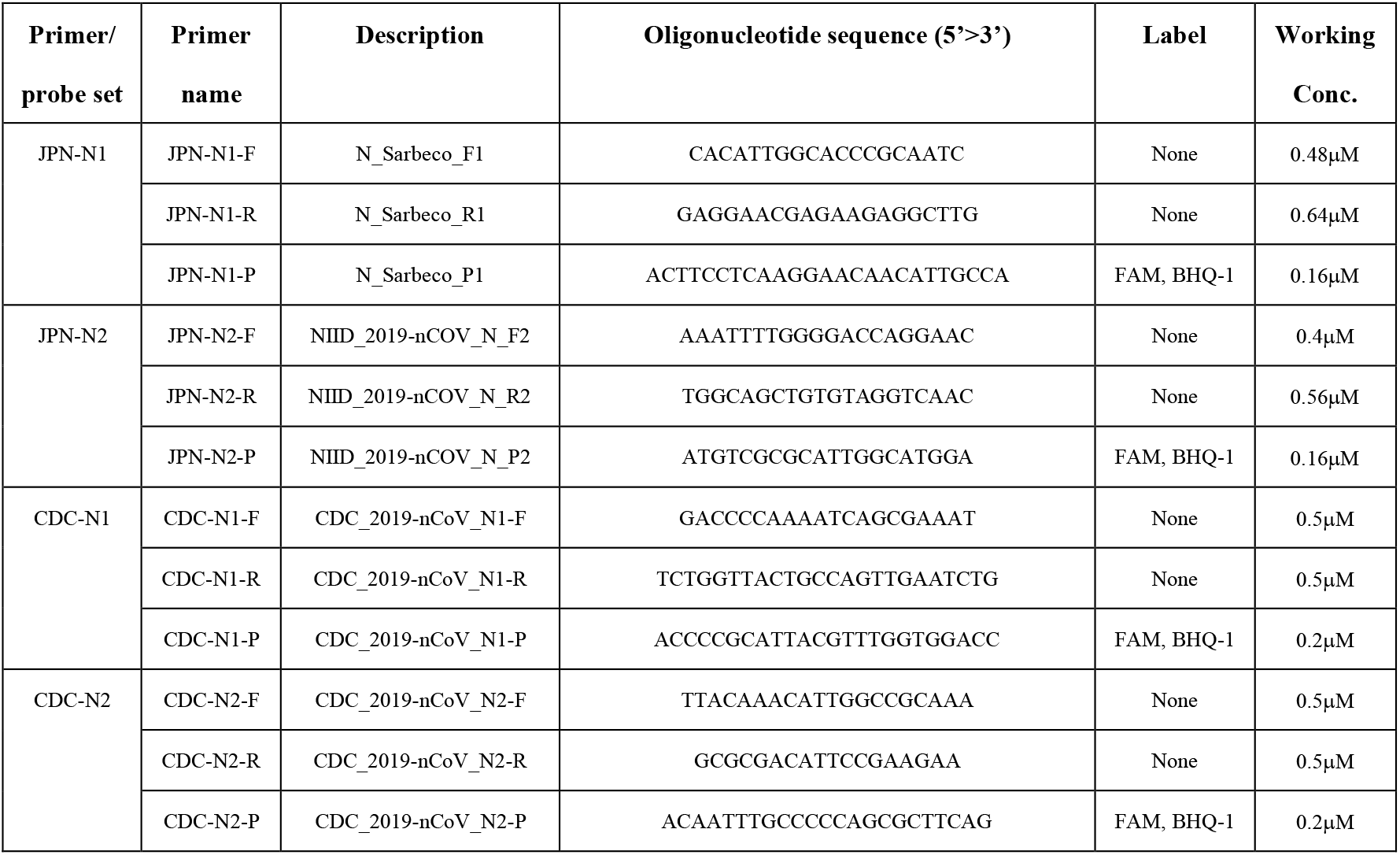
List of primer-probe sets used in this study to check sensitivity and accuracy.

The cycling conditions consisted of RT at 50° Celsius for 5 minutes, initial denaturation at 95° Celsius for 10 sec, and 45 cycles of denaturation at 95° Celsius for 5 seconds and annealing/extension at 60° Celsius for 30 seconds.

## RESULTS

All samples were tested simultaneously for JPN-N2, JPN-N1, CDC-N2, and CDC-N1 primer/probe set (Note: As of May, 2020, the Japanese testing protocol for COVID-19 is only JPN-N2 as announced by the Japan’s NIID).

Table 2 outlines that 85 samples (82 saliva samples and 3 nasopharyngeal swab samples) of a total 102 samples that were tested positive for JPN-N2 primer/probe set while 17 samples (15 saliva and 2 nasopharyngeal swab samples) were tested negative.

**Table 2.**
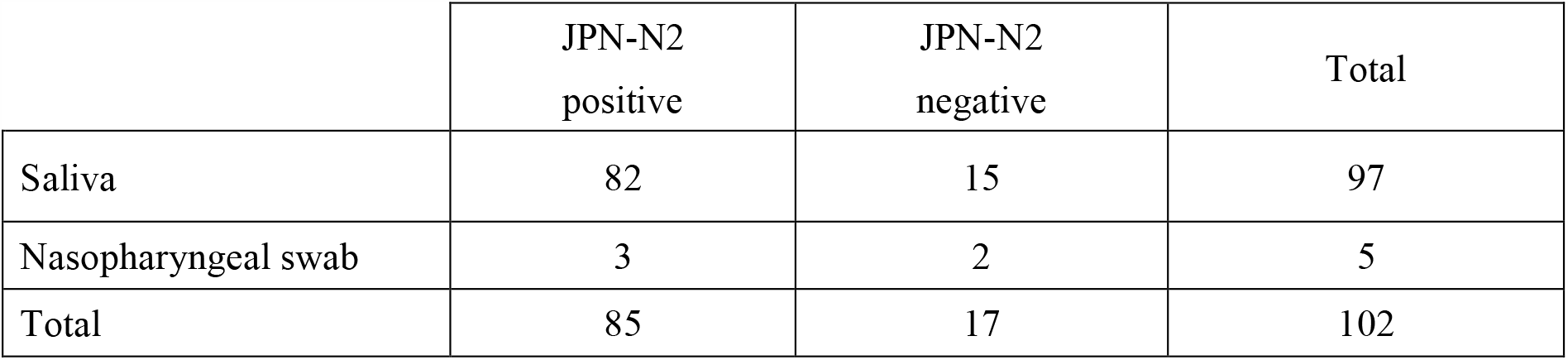
Distribution of Positive SARS-CoV-2 Samples for JPN-N2.

However, Table 3 depicts that among the 17 samples which were tested negative for JPN-N2, 12 samples were tested positive for CDC-N2 primer/probe set while negative for CDC-N1, 4 samples tested positive for both CDC-N1 and CDC-N2. Furthermore, one sample was tested positive for CDC-N1 while negative for CDC-N2 (Figure 3-5).

**Table 3.** Breakdown of JPN-N2 negative samples having positive response from CDC-N1 and/or CDC-N2.

|  | CDC-N1<br>positive | CDC-N1<br>negative | CDC-N1<br>positive | Total |
| --- | --- | --- | --- | --- |
|  | CDC-N2<br>negative | CDC-N2<br>positive | CDC-N2<br>positive |  |
| Saliva | 1 | 10 | 4 | 15 |
| Nasopharyngeal swab | 0 | 2 | 0 | 2 |
| Total | 1 | 12 | 4 | 17 |

**Figure 2.**
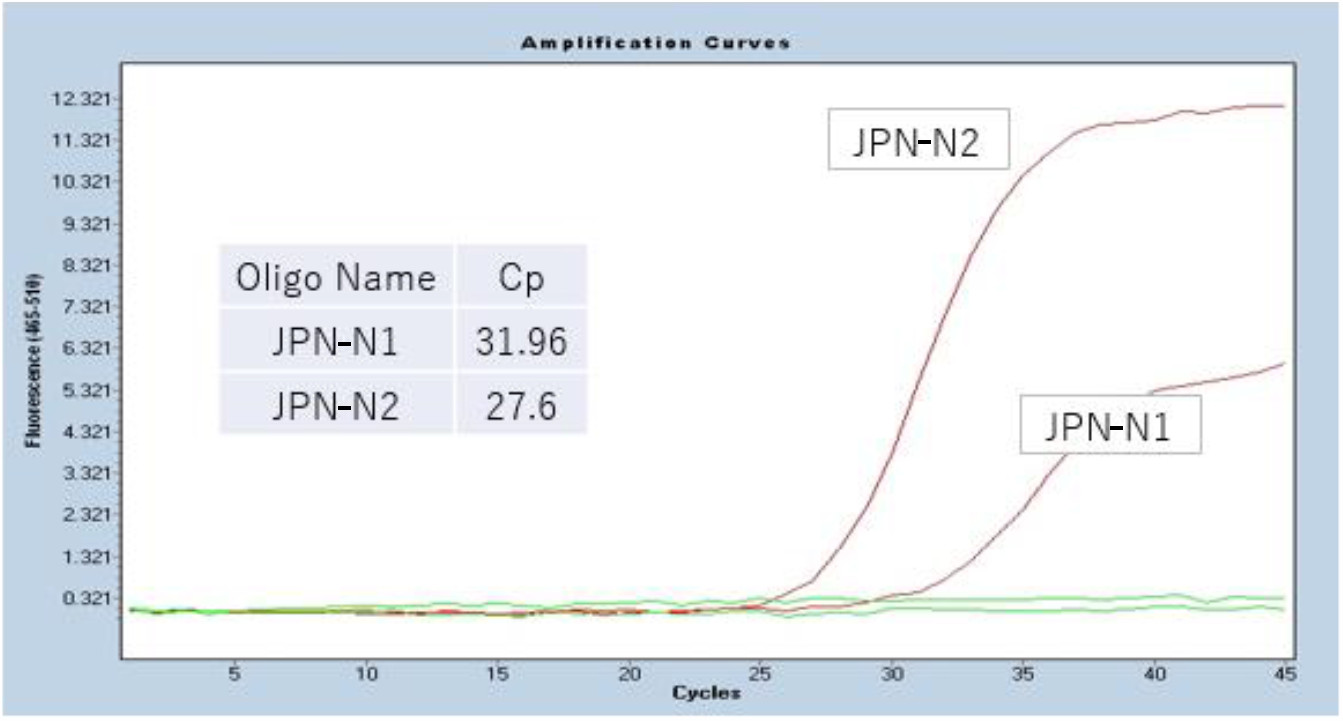
A RT-PCR results for JPN-N2 and JPN-N1 primer-probe sets: JPN-N2 shows relatively higher signal intensity compared to JPN-N1 and the rising curve starts at 25 cycles for SARS-CoV-2 positive samples.

**Figure 3.**
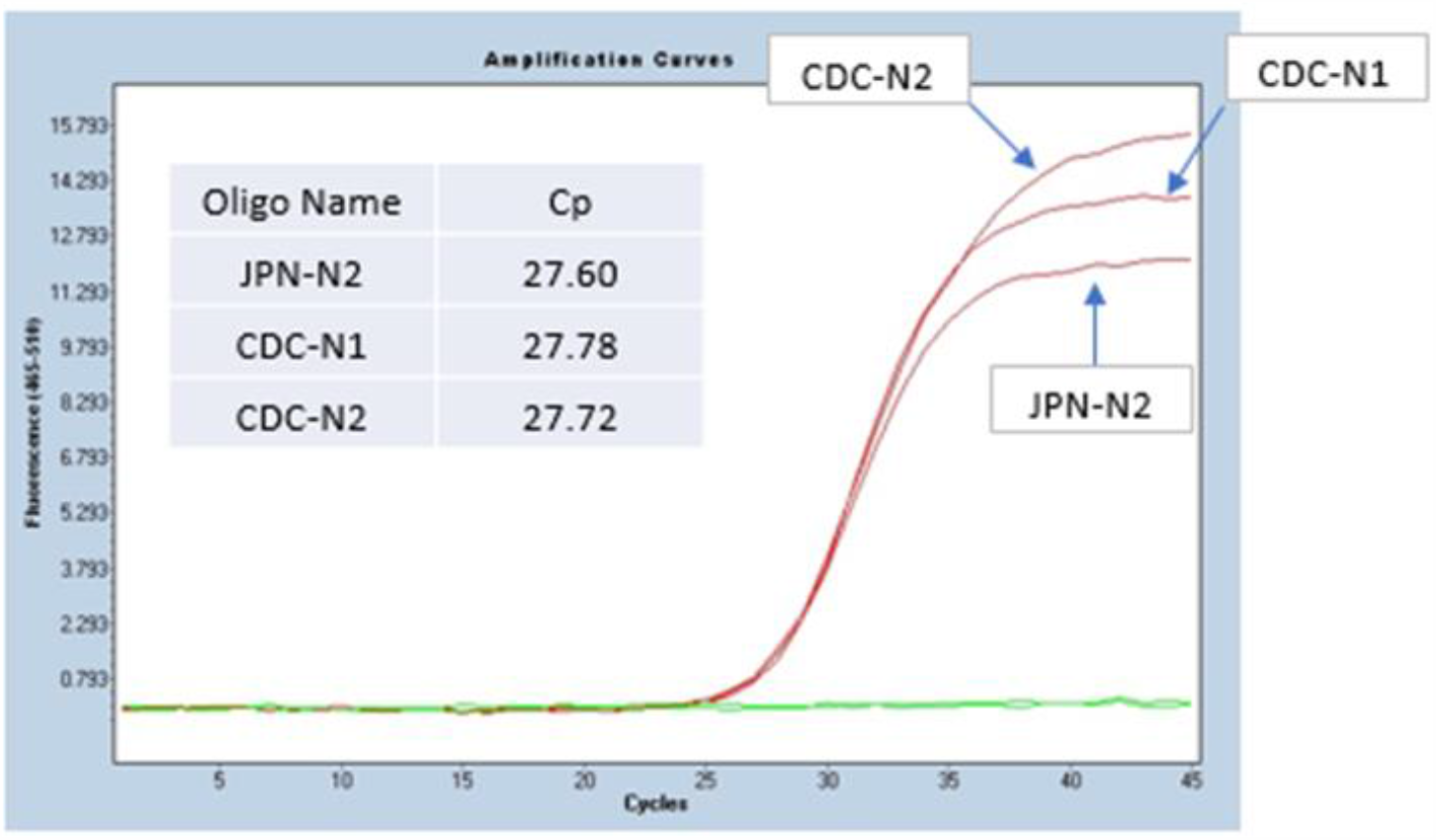
Testing Protocol by Genesis Healthcare incorporating various primers including Japan’s Center of Infectious Disease Testing Protocol (Primers: JPN-N2, JPN-N1, CDC-N2, CDC-N1).

These 17 “*test result irregularities*”, which accounts for 16.7% of total 102 positive sample pool would be declared negative if a laboratory only tested JPN-N2. This high rate of irregular occurrence stemming from 1 primer/probe can be considered as one of the major causes leading to false negatives that are often reported in association with COVID-19 testing accuracy. Furthermore, from the observation above, a combination of multiple primers, in this case, a combination of JPN-N2 and CDC-N2 primer shows the highest rate of accuracy and sensitivity compared to JPN-N2 primer alone, followed by a triple combination of JPN-N-2, CDC-N2 and CDC-N1 for the Japanese population. These combinations, however, may change with various COVID-19 strains that could exist in other populations and further investigation is necessary to identify the optimum combination of primers for the Japanese population as cross-border travel brings different COVID-19 strains that react to different primers.

### Sample Observation of RT-PCR Data of Irregular Samples

Below are several examples of unique and irregular samples and its RT-PCR data that could serve as future hypothesis for identifying and detecting the causes of false testing results.

In the first case, samples reacted only to the CDC-N2 primer but not to the JPN primers. One of the most problematic cases observed are the samples whereby JPN-N2 primer resulted in negative albeit responding positive to CDC-N2 or CDC-N1 primers. Since Japanese testing protocol for COVID-19 is JPN-N2 alone, registered Japanese clinical laboratories are expected to call based on N2 primer result. However, in instances such as these, the importance to provide additional information regarding the test result for CDC is recognized and attending physicians are strongly urged to retest the patient again via RT-PCR or utilize other testing methods such as CT Scan and antigen. This would diminish the risk of misinformation and potentially spread of the infection to others for patients, whether symptomatic or asymptomatic.

Among the 4 primers and probe, only CDC-N2 reacted as positive response at late cycle (40x). This patient (male patient A) became symptomatic 14 days before testing and upon taking PCR test on day 5, tested positive. After 1 week of quarantine and the public health office has lifted his self-quarantine, he voluntarily retested to reconfirm his status. His results showed that his test result would be negative under Japan NIID’s JPN-N2 standalone testing protocol, but the results clearly identified that he is still positive on CDC-N2 and would be declared positive if this person were to be tested at a laboratory in the US, where CDC-N2 is the standard protocol.

The second case also reflects how JPN-N2 was negative while CDC-N1 and CDC-N2 both showed positive responses at later cycles than JPN-N2 positive control (36.54 cycles and 37.44 respectively).

## DISCUSSION

There are numerous factors that could affect RT-PCR result discrepancy, ranging from sample collection method, collection timing, virus inactivation technique, and many other RT-PCR procedures. However, given that the testing protocol strictly follows NIID and CDC testing guideline and protocol, that left viral diversity as a strong candidate for the irregularities.

It’s hypothesized that various virus strains align differently to different primers used globally due to its unique sequence. Specifically, at the time of analysis, there are 15,745 SARS-CoV-2 sequences submitted to NCBI GenBank (Sayers et al. 2020) where each sequence contains different combination of variants/mutations. Therefore, to further validate our assumption, the virus mutation data and primers oligonucleotide sequences are investigated.

### Primers vs. Viral Genetic Variations

In an effort to try to explore the primer(s) effectiveness in current virus variations, an exploratory data analysis has been conducted over 84 virus strains from Japan, retrieved from NCBI GenBank genetic sequence database as of August 28, 2020.

From extracted Japan’s samples, a phylogeny tree was generated using tool from NCBI Virus (Hatcher et al. 2017). The resulting tree shows different clades connected in hierarchies, suggesting that there are different virus mutations in the geographical region. Next, a sequence alignment using primers nucleotide from different countries against all coronavirus 2 strains from Japan was performed, using Blastn. The alignment results are then filtered to contain only perfect consecutive alignment of all nucleotides, meaning identities must be exactly equal to the length of primer oligonucleotide sequence with no gap. With such a rigid filter, the number of alignment matches were visualized using iTOL (Letunic and Bork 2019) and many virus strains those do not match with some primers are found as shown in Figure 10.

**Figure 4.**
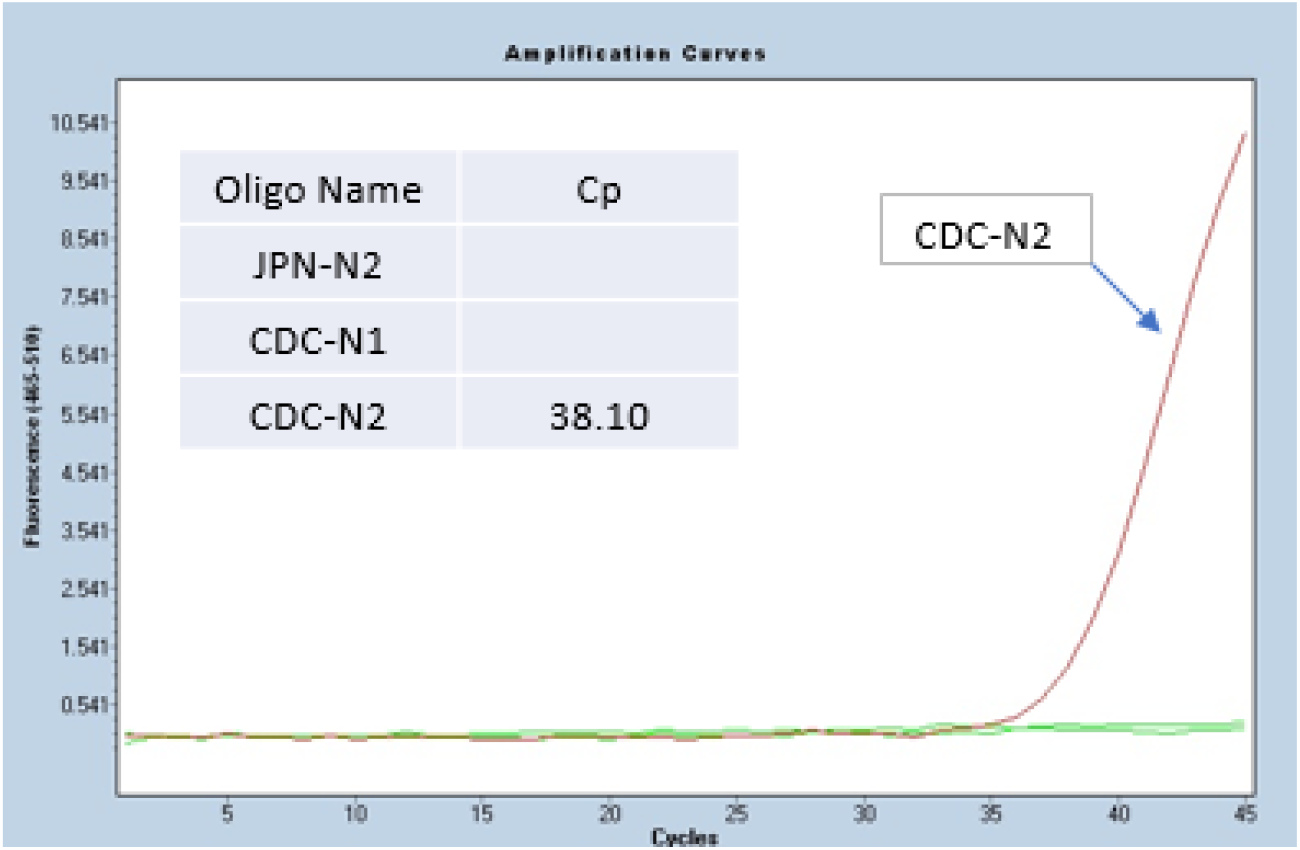
A Japanese sample that was negative for JPN-N2 and CDC-N1 but positive for CDC-N2.

**Figure 5.**
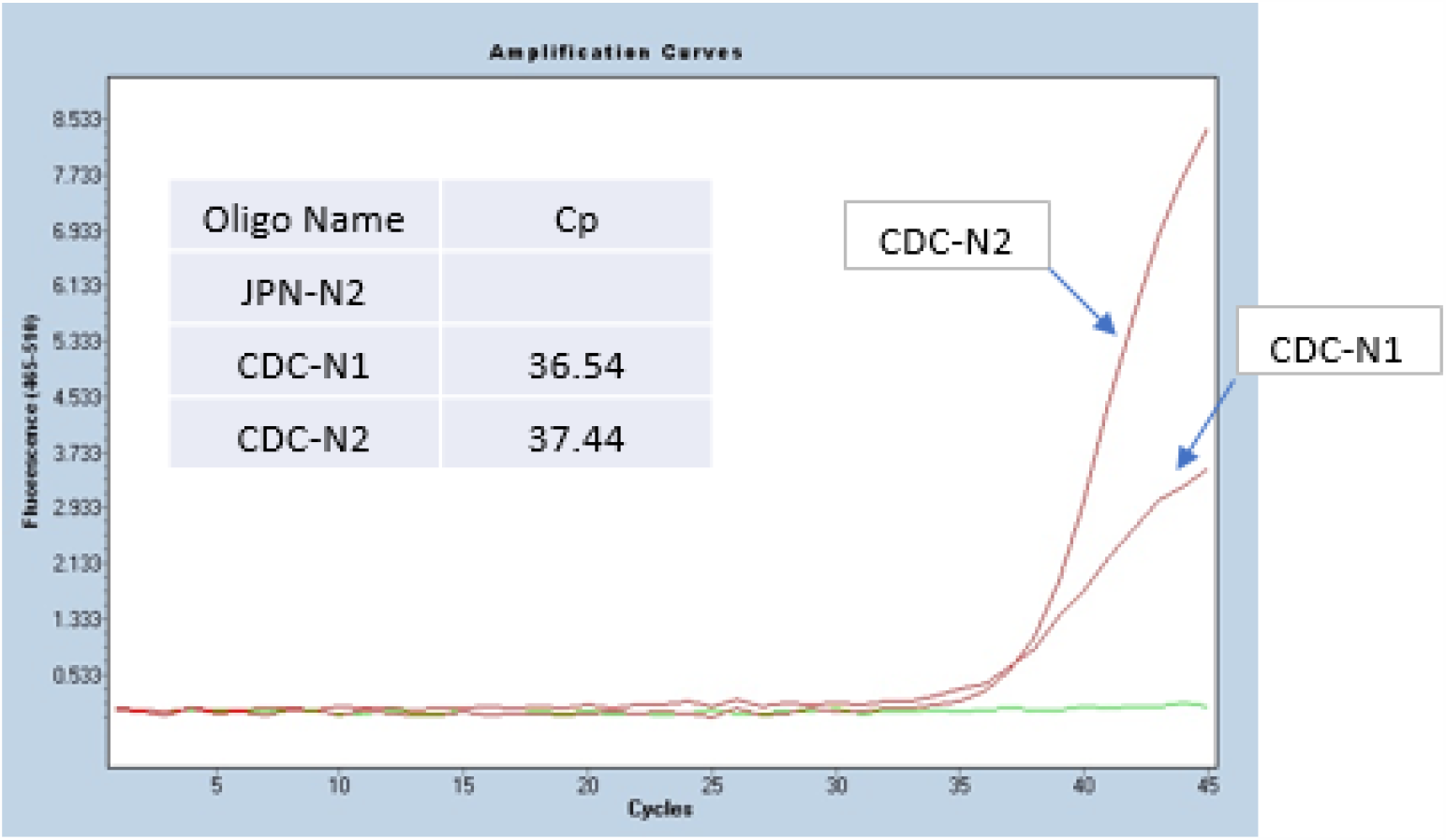
An example of a Japanese sample that was negative for JPN-N2 but showed positive to both CDC-N1 and CDC-N2 primer-probe sets.

**Figure 6.**
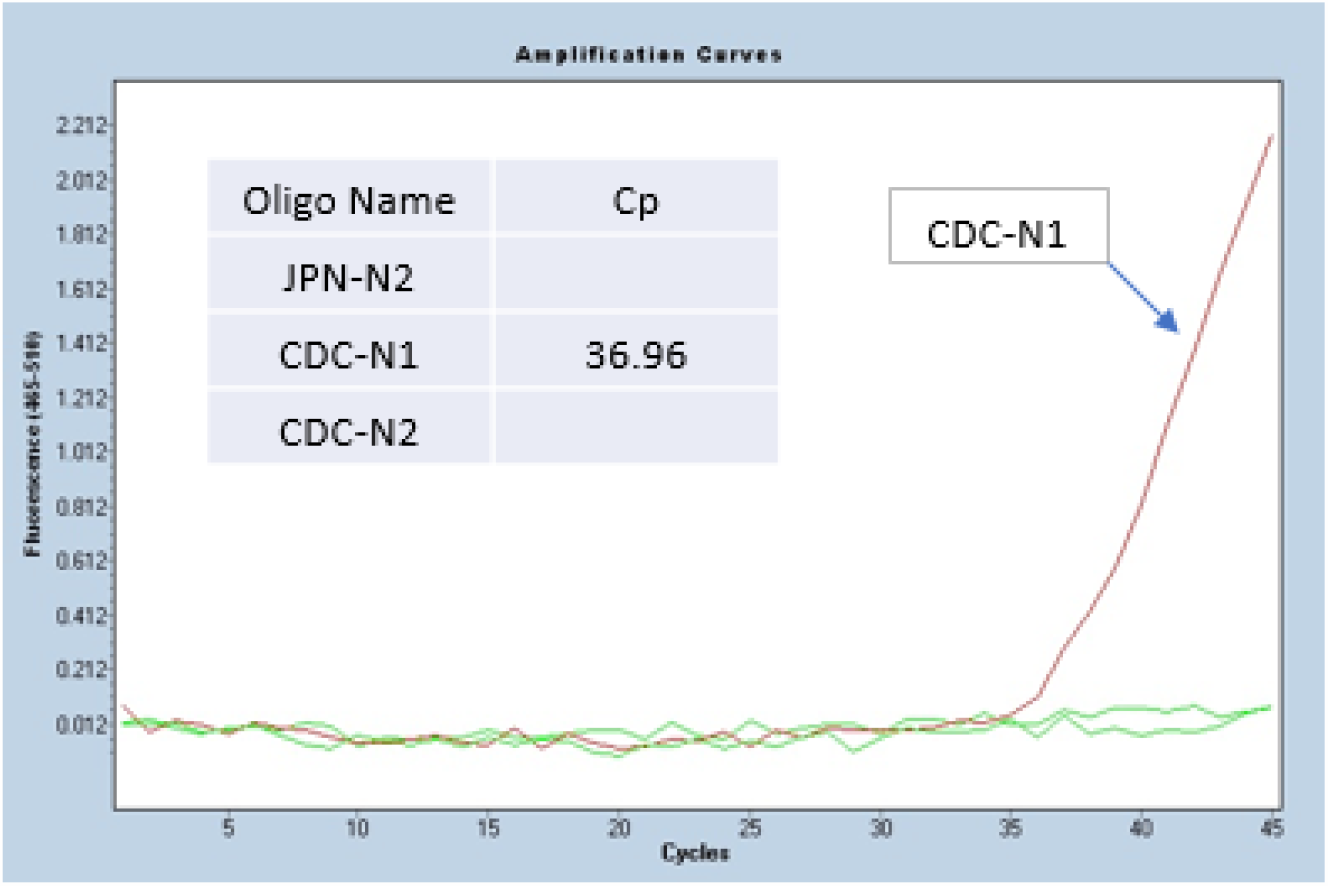
A Japanese sample that was negative for JPN-N2 and CDC-N2 while positive for CDC-N1 primer-probe sets.

**Figure 7.**
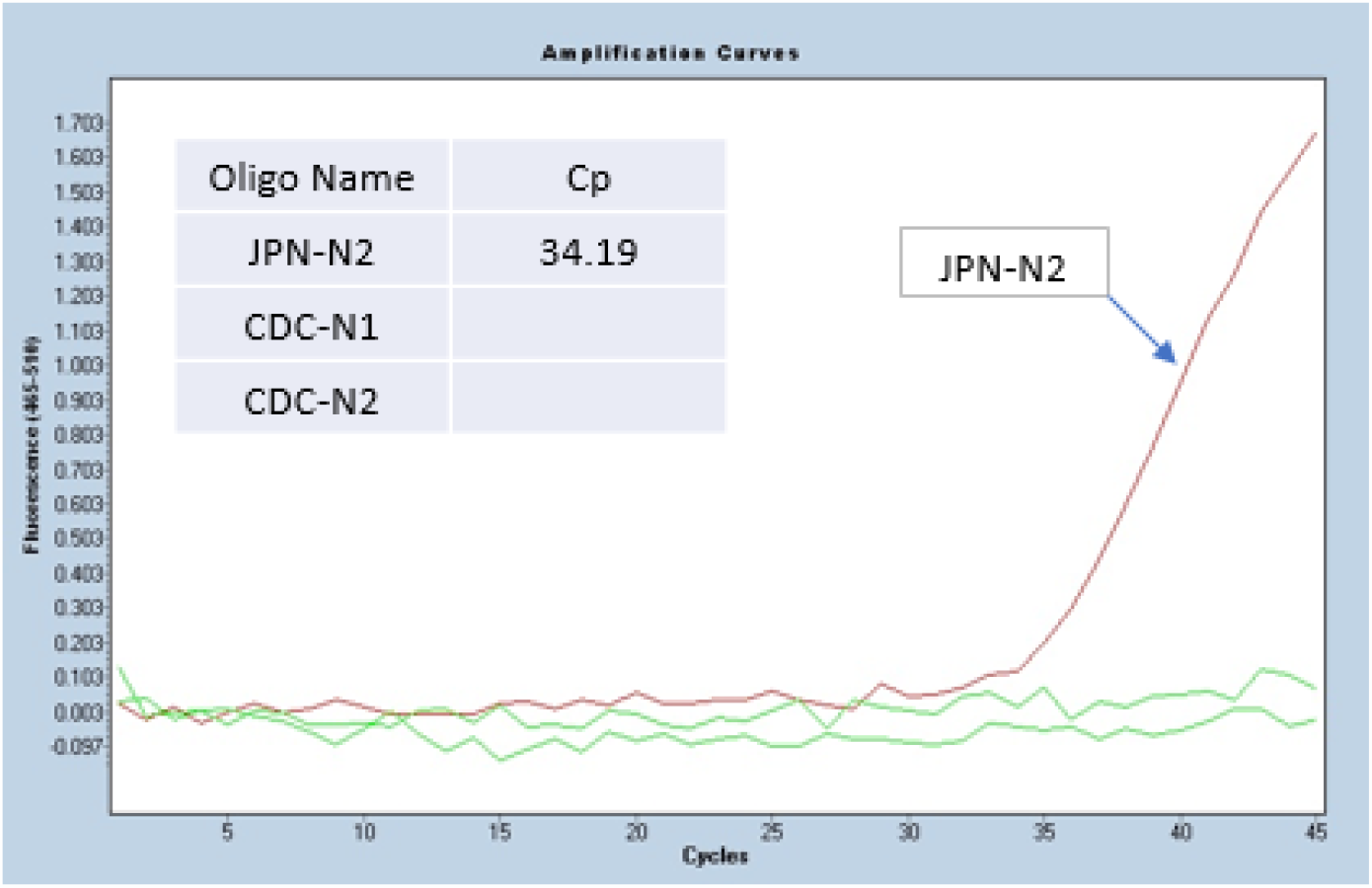
A Japanese sample that was positive for JPN-N2 however was negative for CDC-N2, CDC-N1 primer-probe sets.

**Figure 8.**
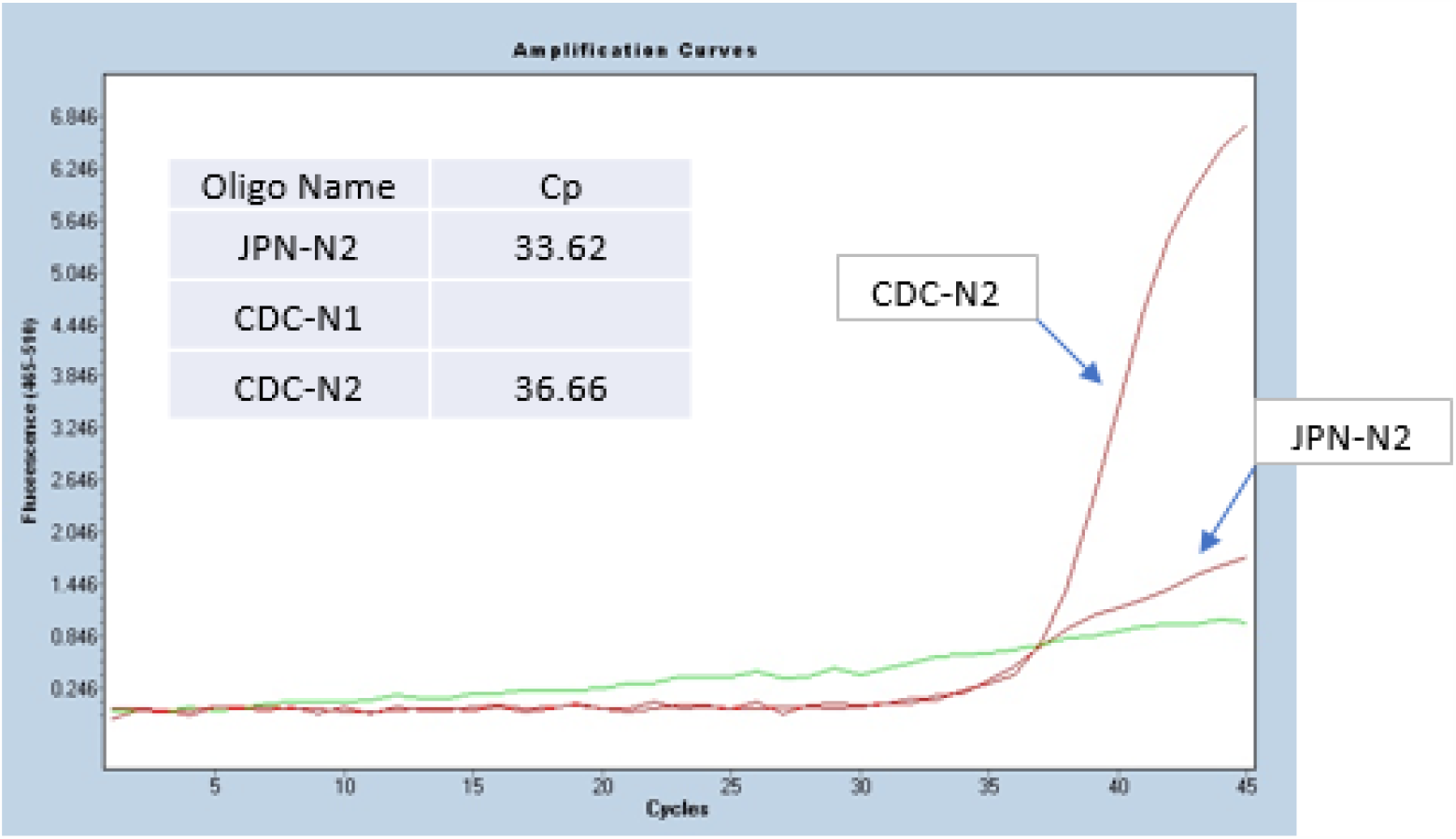
A Japanese sample that was positive for CDC-N2 but weakly positive for JPN-N2 and negative for CDC-N1 primer.

**Figure 9.**
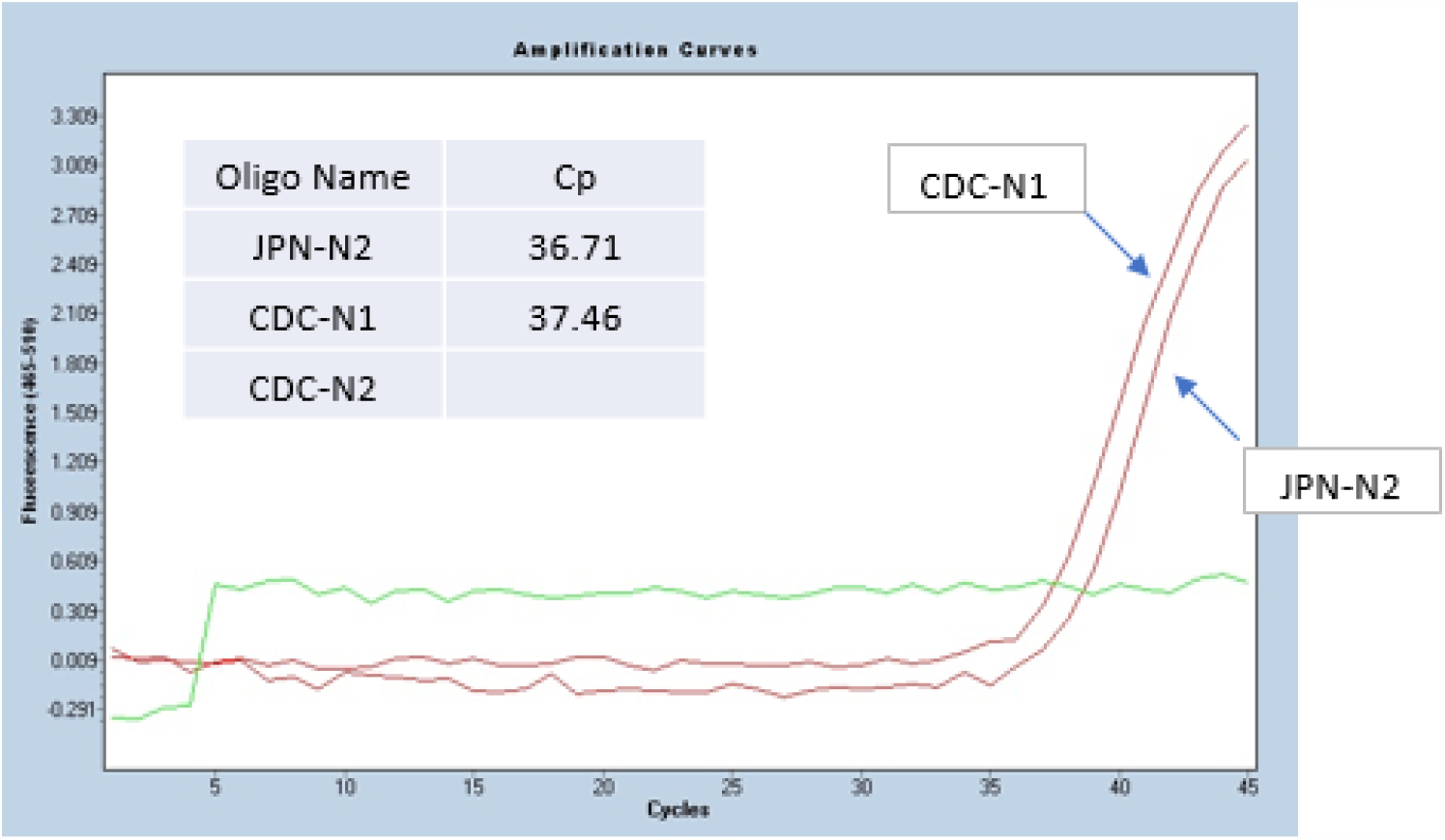
A Japanese sample that was positive for both JPN-N2 and CDC-N1 but negative for CDC-N2 primer.

**Figure 10.**
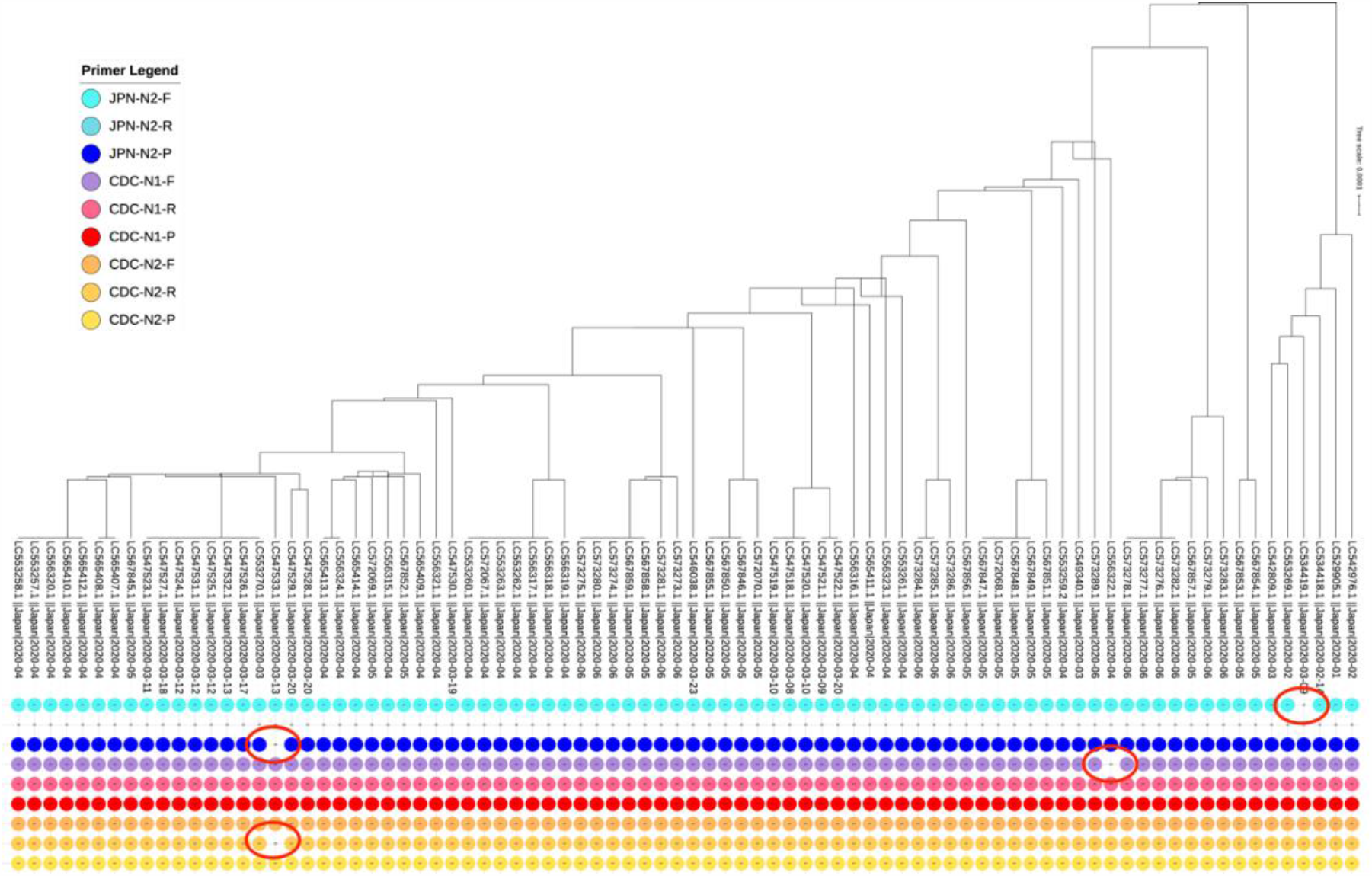
84 SARS-CoV-2 sequences identified for Japanese COVID samples with primers locations where false negatives will most likely occur.

The finding also coincides with other study (Osório and Correia-Neves 2020) reporting that of 33 oligonucleotides developed by different centers and shared by WHO, 79% (26) has at least one genome mutation at the primer binding sites from GISAID database containing 1,825 SARS-CoV-2 genomes.

While one dominant strain will test positive for a specific primer, there are irregularities that will not display positive for another primer, most likely resulting in false negative calls. In this respect, laboratories should examine their primers against all known viral genome sequences to ensure that the selected primers will not result in inaccurate calling.

## CONCLUSION

The conflicting positive/negative results from the same samples that occur among different primers (JPN-N2, JPN-N1, CDC-N2, CDC-N1) outline 1) the possibility of high occurrences of false negative testing results by laboratories where JPN-N2 is the only primer used for SARS-CoV-2, and 2) the probability that false negative/positive will likely occur when the primer binding site lies in the virus mutated location for JPN-N2 and CDC-N2.

This initial observation also suggests testing CDC-N2 in addition to JPN-2 primer, especially for symptomatic patients, with the objective to reduce false negatives and increase accuracy. Lastly, the inclusivity of CDC-N1 will result further in reducing false negatives but using CDC-N1 alone may also result in false positive calling and therefore should not be used without either JPN-N2 or CDC-N2.

Further investigation by sequencing these irregular samples will identify which primers react to different strains of the virus existing among different populations. Secondly, these irregularities raise the need for the international testing community to explore a standardized universal primer-probe sets combination for more accurate RT-PCR-based COVID-19 testing, which shall help to detect different virus strain’s reaction to selected primers by each country’s testing protocol. This is a critical consideration to ensure accurate diagnostic testing results entailing public health and safety. While more countries are re-opening their borders and resuming international travel to sustain the economic growth, this may also lead to the introduction of new strains to different populations furthering the demand for SUP.

## Data Availability

The data used and produced in this research are available at: https://github.com/genesis-healthcare/covid-19_pcr_irregularities　

## Acknowledgements

We acknowledge the physicians from the originating medical facilities responsible for obtaining the specimen from patients.

## Conflict of Interests

The authors declare that there is no conflict of interest.

## Ethics Statement

Biological samples were collected from volunteers, self-medication testing participants and patients under physician care administering clinical testing for SARS-CoV-2 (COVID-19). Samples were anonymized and de-identified, so laboratory technicians and researchers were blind to the identity of the patients. Only secondary non-identifying data including age, biological sex and symptoms were provided. All experimental protocols were approved by Genesis Healthcare’s Ethics Committee.

## Funding Sources

This work is supported by Genesis Healthcare Corporation. The sponsor has no influence in the study other than providing resources and facility in conducting research and publication.

## Notes

### Competing Interest Statement

The authors have declared no competing interest.

### Clinical Protocols

https://www.niid.go.jp/niid/images/epi/corona/2019-nCoVmanual20200217-en.pdf

### Summary of Updates

Revert a missing author.

